# Association between physicians’ maldistribution and core clinical competency in resident physicians

**DOI:** 10.1101/2023.10.26.23297546

**Authors:** Kiyoshi Shikino, Yuji Nishizaki, Koshi Kataoka, Masanori Nojima, Taro Shimizu, Yu Yamamoto, Sho Fukui, Kazuya Nagasaki, Daiki Yokokawa, Hiroyuki Kobayashi, Yasuharu Tokuda

**Affiliations:** Department of Community-Oriented Medical Education, Chiba University Graduate School of Medicine, Chiba, Japan; Department of General Medicine, Chiba University Hospital, Chiba, Japan; Division of Medical Education, Juntendo University School of Medicine, Tokyo, Japan; Center for Translational Research, The Institute of Medical Science, The University of Tokyo, Tokyo, Japan; Department of Diagnostic and Generalist Medicine, Dokkyo Medical University Hospital, Tochigi, Japan; Division of General Medicine, Center for Community Medicine, Jichi Medical University, Tochigi, Japan; General Medicine, Kyorin University School of Medicine, Tokyo, Japan; University of Tsukuba, Tsukuba, Ibaraki, Japan; Department of Internal Medicine, Mito Kyodo General Hospital, Tsukuba, Ibaraki, Japan; Department of Medical Education, Kyushu University, Fukuoka, Japan; Muribushi Okinawa Center for Teaching Hospitals, Okinawa, Japan

**Keywords:** clinical core competency, general medicine in-training examination, physician maldistribution, physician uneven distribution index, resident physician

## Abstract

**Importance:** This study highlights the association between physicians’ maldistribution and core clinical competency of resident physicians and emphasizes the global significance of addressing healthcare access disparities.

**Objective:** To investigate the relationship between a prefectural program with and without physician maldistribution and core clinical competency, measured using the General Medicine In-Training Examination (GM-ITE).

**Design:** Cross-sectional study.

**Setting:** Data from the GM-ITE survey were collected in January 2023.

**Participants:** Resident physicians in their first and second postgraduate year (PYG-1 and PGY-2) who were employed at Japanese hospitals that required the GM-ITE or resident physicians who voluntarily participated in the GM-ITE.

**Exposure:** Physician uneven distribution (PUD) index is a policy index developed and adopted in Japan. It serves as an indicator of regional disparities among physicians within the country. A low PUD index indicates that there is an insufficient medical supply relative to the medical demand in that region.

**Main Outcomes and Measures:** The GM-ITE scores of resident physicians.

**Results:** The high PUD index group included 2,143 participants and the low PUD index group included 1,580 participants. After adjusting for relevant confounders, multivariable linear regression analyses revealed that the low PUD index group had a significantly higher GM-ITE score compared to the high PUD index group (adjusted coefficient: 1.14; 95% confidence interval: 0.62–1.65; p<0.001).

**Conclusions and Relevance:** Resident physicians in regions with low PUD indices had significantly higher GM-ITE scores. These findings underscore the significance of addressing physician maldistribution to enhance the clinical competency of resident physicians and emphasize the potential benefits of reducing regional healthcare disparities, particularly in terms of medical education and training. These insights have broader relevance for healthcare policies and medical training programs worldwide, highlighting the need to consider physician distribution as a critical factor in improving healthcare access and quality.

**KEY POINTS:** *Question:* How does physician maldistribution, indicated by the physician uneven distribution (PUD) index, impact resident physicians’ clinical competence based on performance in the General Medicine In-Training Examination (GM-ITE) in Japan?

*Findings:* In this nationwide cross-sectional study, resident physicians affiliated with hospitals in regions with a lower PUD index (indicating insufficient medical professional supply relative to healthcare demands) had significantly higher scores on the GM-ITE than those in regions with a higher PUD index.

*Meaning:* Training resident physicians in areas short of physicians does not adversely affect their education; rather, it can enhance medical education and address the physician maldistribution issue.

## INTRODUCTION

Japan is currently experiencing a severe crisis in local healthcare delivery due to the uneven distribution of physicians, which is exacerbating the critical situation in community hospitals in rural areas.^1,2^ The insufficient number of physicians in rural areas due to urbanization remains a serious problem.^3–5^ The issue of physician maldistribution is not unique to Japan; it is a global concern affecting China, India, England, Canada, the United States, Australia, and European nations.^6–11^

In 2019, the Ministry of Health, Labour and Welfare (MHLW) initiated an intervention policy aimed at rectifying geographical disparities in physician distribution at the prefectural level.^1,12–14^ This policy introduced the physician uneven distribution (PUD) index. The PUD index was designed to gauge the extent of physician maldistribution by considering both medical supply and demand at the prefectural level (Supplement 1). A lower PUD index indicates a higher level of uneven distribution of physicians across prefectures.

In Japan, training physicians to produce proficient clinical practitioners involves 6 years of medical school followed by 2 years of clinical training as resident physicians.^15–17^ Resident physicians are required to undergo supervised training and rotate through seven specialties (internal medicine, surgery, pediatrics, obstetrics and gynecology, psychiatry, emergency medicine, and community-based medicine). During this 2-year clinical training period, resident physicians are mandated to acquire core clinical competency that enables them to effectively manage common medical conditions in general practice, irrespective of their future specialization.^18^ This clinical training system is currently undergoing a policy review, addressing concerns regarding recruitment capacity and other aspects, with a view to mitigate physician maldistribution in various regions and enhance the quality of clinical training.

The MHLW has conducted a questionnaire regarding clinical training in areas characterized by an uneven distribution of physicians.^19^ According to the report, resident physicians who received clinical training in areas with a relatively low number of physicians reported a higher level of satisfaction.^19^ Of the 2,684 resident physicians who received their clinical training in areas with a relatively high number of physicians, 74.2% expressed satisfaction, though 79.6% of the 1,703 resident physicians who received their clinical training in areas with a relatively low number of physicians reported satisfaction with their clinical training.^19^ A questionnaire assessing self-perceived competencies related to clinical training revealed no significant differences between prefectures with a relatively high number of physicians and those with a low number of physicians in terms of professionalism (69.4% vs. 69.4%), medical knowledge and problem-solving skills (69.9% vs. 67.2%), medical skills and patient care (71.6% vs. 70.1%), or quality and safety of care (68.2% vs. 68.0%).^19^

However, to the best of our knowledge, no objective and quantitative assessment of clinical training and proficiency in core clinical competencies within regions affected by physician maldistribution has been conducted in Japan.^19,20^ The primary objective of this study was therefore to evaluate the relationship between areas with a relatively high number of physicians and areas with a relatively low number of physicians in each prefectural program and the achievement of core clinical competencies, measured using resident physicians’ General Medicine In-Training Examination (GM-ITE) scores.^21^

## METHODS

### Study design, setting, and participants

A nationwide cross-sectional survey was conducted on 9,011 postgraduate resident physicians (approximately 50% of the total number of resident physicians in Japan) from 662 hospitals who had taken the GM-ITE between January 17–30, 2023. The participants in this study included resident physicians in their first and second postgraduate years (PYG-1 and PGY-2, respectively) in Japanese hospitals in which the GM-ITE was included in the residency program as well as those who voluntarily participated in the GM-ITE. We included the participants of the GM-ITE 2022. Participants who refused the use of their data for research were excluded.

### Main outcome

The main outcome of this study was the total GM-ITE scores of resident physicians. In 2011, the nonprofit Japan Institute for Advancement of Medical Education Program (JAMEP) developed the GM-ITE as an in-training examination of clinical knowledge using a methodology similar to that of the US Residency Internal Medicine In-Training Examination.^21–23^ The GM-ITE provides resident physicians and program directors with an objective, reliable, and valid assessment of core clinical competency, including clinical knowledge.^21–24^ Designed by a committee of experienced attending physicians organized by the JAMEP, the 2-hour GM-ITE includes 80 multiple-choice questions in several domains^24,25^ and is scored from 0 to 80, with higher scores indicating better performance and knowledge of clinical medicine.

An assessment of the validity of the GM-ITE revealed a strong positive correlation between GM-ITE scores and scores on the Professional and Linguistic Assessments Board test, Part 1 (PLAB 1), which was designed to assess the depth of medical knowledge and levels of medical and communication skills.^26^ In validity testing, the discrimination index indicates how well the item differentiates between students with high and low aptitude (whether high-aptitude learners performed better, worse, or the same as that of low-aptitude learners).^27^ The GM-ITE exhibited equivalent or superior discriminative power compared to the PLAB 1 examination.^26^

### Exposure and covariates

The exposure in this analysis was the PUD index (updated on August 9, 2023). This index is a policy index developed and adopted in Japan. It serves as an indicator of regional disparities among physicians within the country. A low PUD index indicates that there is an insufficient medical supply relative to the medical demand in that region. Resident physicians were categorized into three groups based on the score ranges of their residency program, as follows: the high PUD index group (PUD index: 353.9-266.9), moderate PUD index group (PUD index: 266.5-230.5), and low PUD index group (PUD index: 228.0-182.5).^19^ The resident physicians were also categorized based on their duty hours per week: <60 hours, 60–79 hours, and ≥80 hours. The duty hour category was defined with a lower limit indicating no or minimal overtime and an upper limit of 80 hours per week, in accordance with the new resident physician duty hour limit set by the MHLW. The questionnaire administered in this study included questions regarding the participants’ demographic information, such as hospital type (community hospital or university hospital), gender, postgraduate year (PGY-1 or PGY-2), participation in the general medicine rotation, and the duration of the participant’s internal medicine rotation. The number of night shifts per month, average number of assigned inpatients, and self-study time per day were also reported by the resident physicians.

### Statistical analysis

The participant demographics and characteristics, as well as the GM-ITE scores, were compared among the three PUD index groups. Multivariate linear regression analysis was used to evaluate the association between the PUD index grade and the GM-ITE score. The results were adjusted for covariates, including hospital type, gender, PGY, rotation in the general medicine department, internal medicine rotation duration, number of night shift duties per month, average number of assigned inpatients, self-study time, and duty hours per week. All statistical analyses were conducted using Statistical Analysis System version 9.4 for Windows (SAS Institute Inc., Cary, NC, USA), with the level of significance set at p < 0.05. All statistical tests were two-tailed.

This study was approved by the Ethical Review Committee of the Japan Organization for Advancing Medical Education (approval no. 23–9). All participants provided informed consent prior to their participation in the study in accordance with the Declaration of Helsinki.

## RESULTS

### Demographics

A total of 5,762 resident physicians from 608 hospitals, including 3,922 (68.1%) men, participated in this study (Table 1). The minority of resident physicians (n=979; 17.0%) were from university hospitals. The high PUD index group included 2,143 participants, the moderate PUD index group included 2,039 participants, and the low PUD index group included 1,580 participants.

**Table 1.**
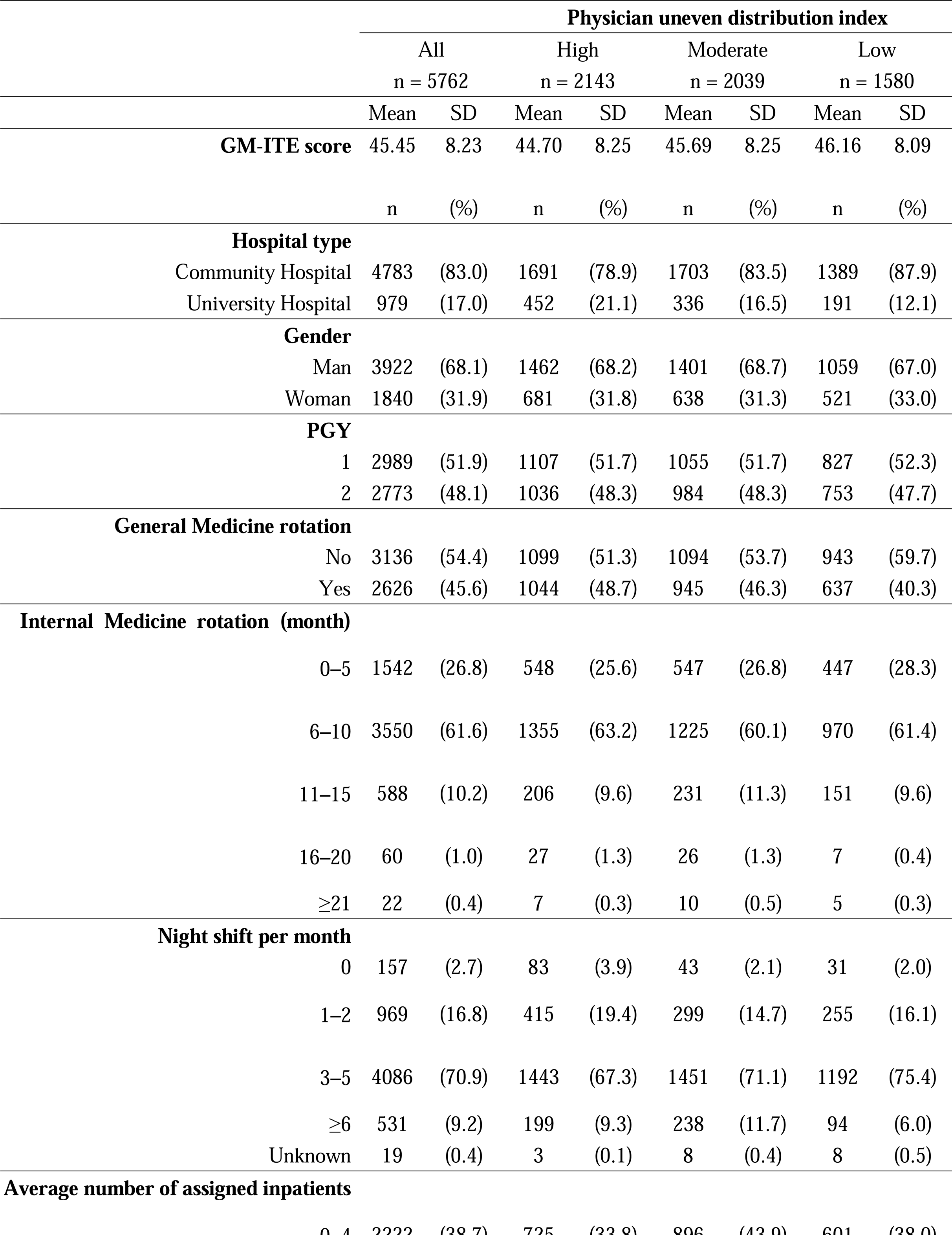
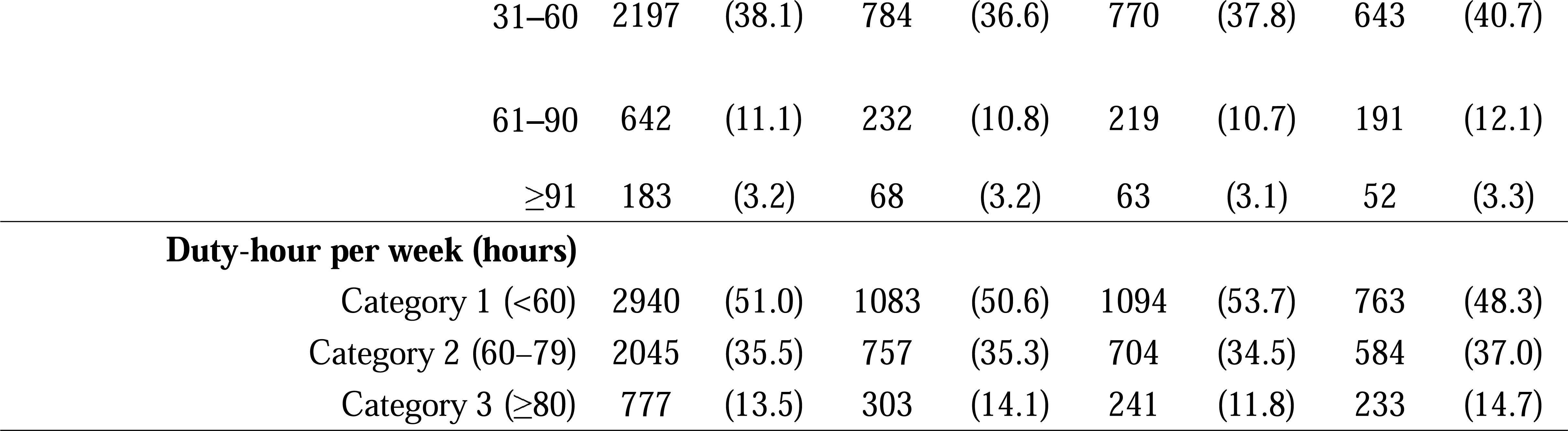
Baseline characteristics of resident physicians categorized according to PUD index. PUD, Physician uneven distribution index

Overall, the mean GM-ITE score was 45.45 ± 8.23 points. The mean scores of the high, moderate, and low PUD index groups were 44.70 ± 8.25 points, 45.69 ± 8.25 points, and 46.16 ± 8.09 points, respectively. The GM-ITE score distribution is shown in terms of the PUD index in Figure 1.

**Figure 1.**
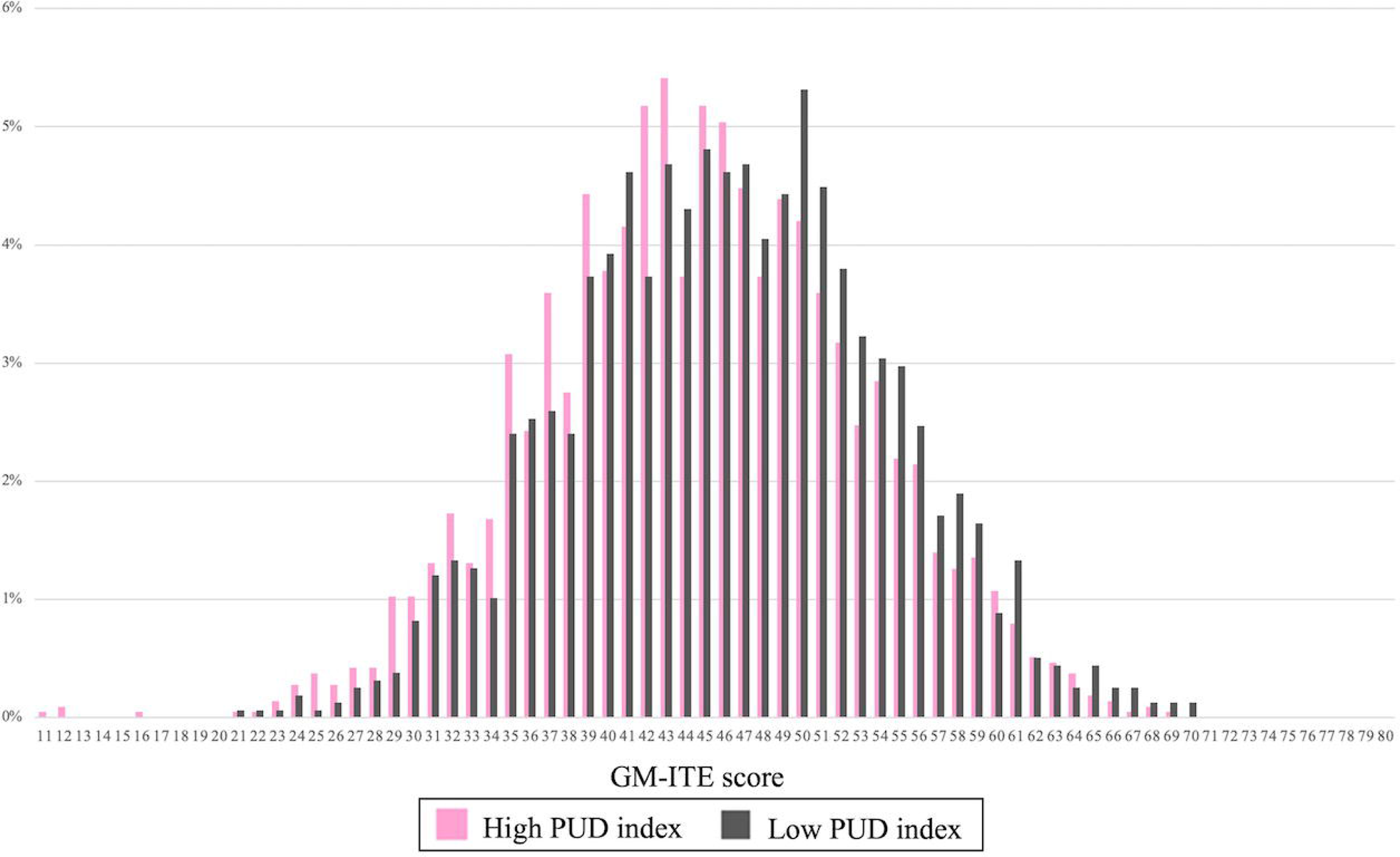
GM-ITE score distribution between low and high PUD index prefecture resident physicians. GM-ITE, General Medicine In-Training Examination

Approximately half (54.4%) of the participants did not undergo a general medicine rotation; 26.8% underwent an internal medicine rotation that lasted 0–5 months, while 61.6% underwent an internal medicine rotation that lasted 6–10 months. The reported hours per week were <60 hours in 51.0% of participants, 60–79 hours in 35.5% of participants, and ≥80 hours in 13.5% of participants.

### Univariate linear regression analysis

As shown in Table 2, in univariate linear regression analysis, a low PUD index was associated with a higher GM-ITE score when compared to a high PUD index (adjusted coefficient: 1.46; 95% confidence interval: 0.93–1.99; p<.001). A moderate PUD index was also associated with a higher GM-ITE score (adjusted coefficient: 0.98; 95% confidence interval: 0.49-1.48; p<.001) when compared to a high PUD index.

**Table 2.**
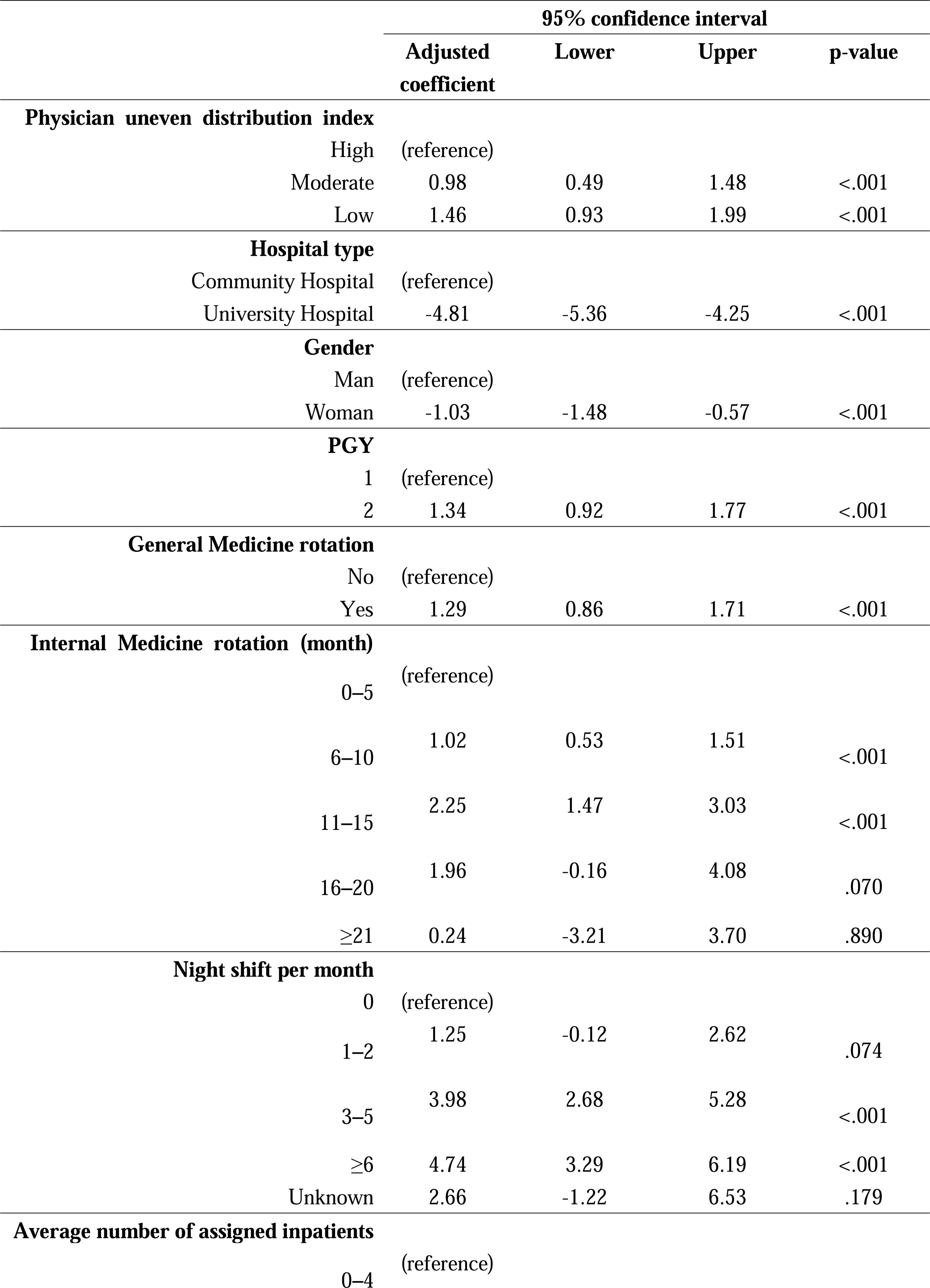
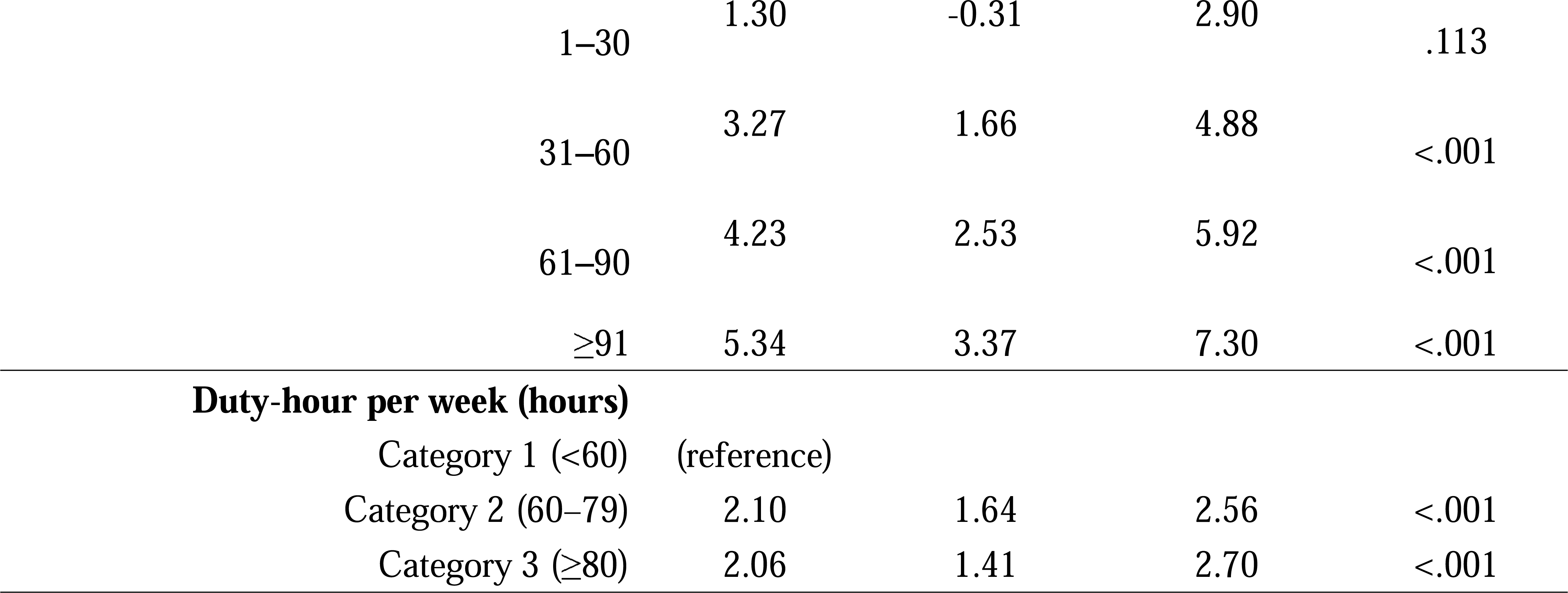
Factors associated with the GM-ITE score in univariate regression analysis. GM-ITE, General Medicine In-Training Examination

### Multivariate linear regression analysis

In multivariate linear regression analysis, a low PUD index was identified as an independent predictor of a higher GM-ITE score when compared to a high PUD index (adjusted coefficient: 1.14; 95% confidence interval: 0.62–1.65; p<.001) (Table 3). A moderate PUD index was also identified as an independent predictor of a higher GM-ITE score (adjusted coefficient: 0.91; 95% confidence interval: 0.43-1.39; p<.001) when compared to a high PUD index.

**Table 3.**
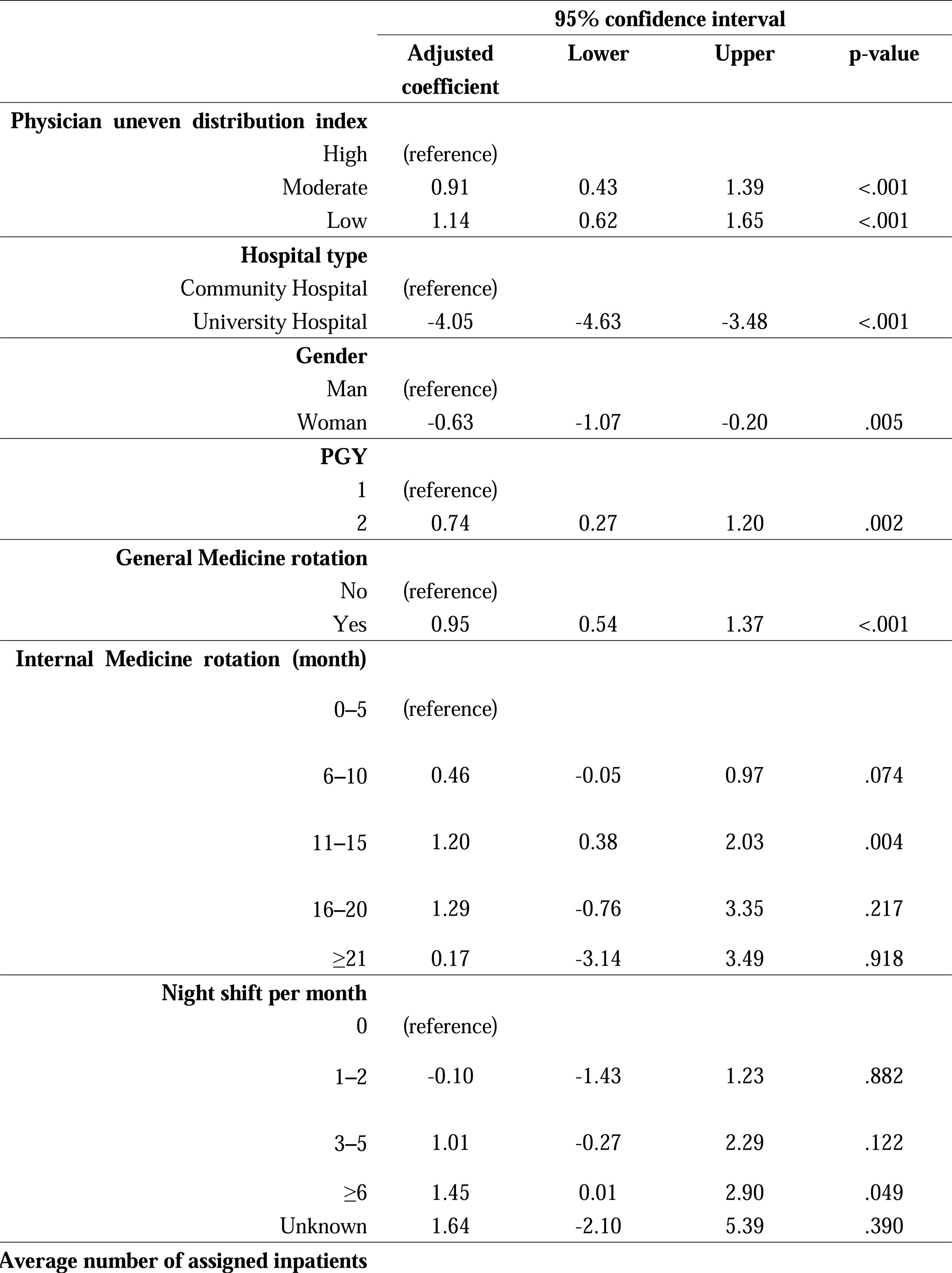
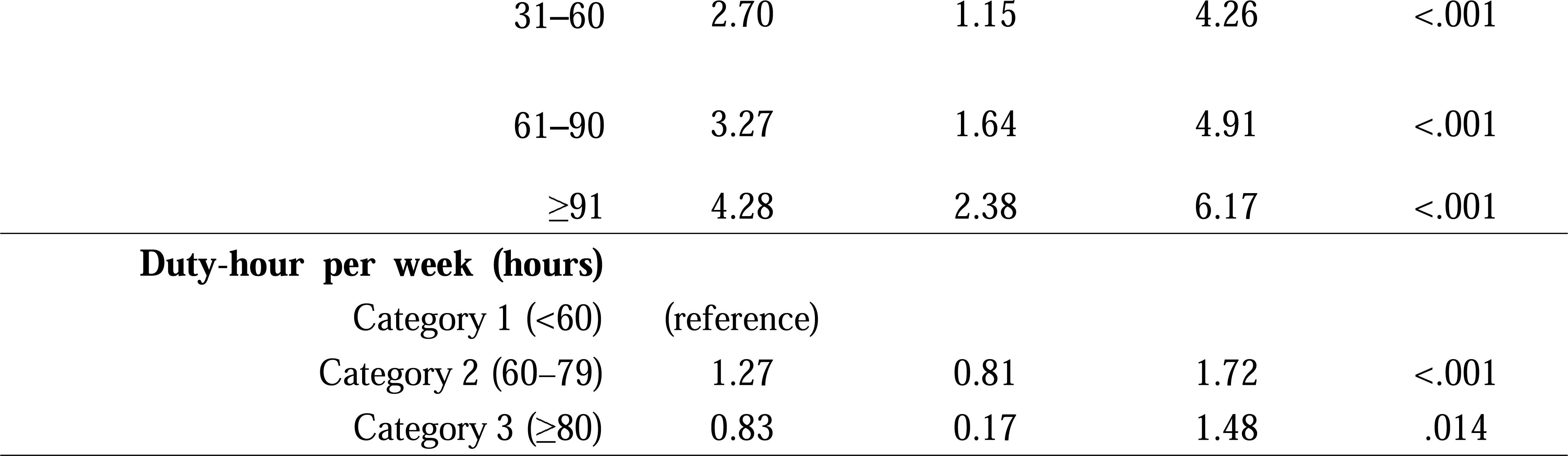
Factors associated with the GM-ITE score in multivariate linear regression analysis. GM-ITE, General Medicine In-Training Examination

Participation in a residency program at a university hospital was identified as an independent predictor of a lower GM-ITE score when compared to participation at a community hospital (adjusted coefficient: −4.05; 95% confidence interval: −4.63--3.48; p<.001). A PGY-2 status was an independent predictor of a higher GM-ITE score (adjusted coefficient: 0.74; 95% confidence interval: 0.27-1.20; p=.002). Having 31–60 (adjusted coefficient: 2.70; 95% confidence interval: 1.15-4.26; p<.001), 61–90 (adjusted coefficient: 3.27; 95% confidence interval: 1.64-4.91; p<.001), or ≥91 (adjusted coefficient: 4.28; 95% confidence interval: 2.38-6.17; p<.001) minutes of study time per week was also indicative of higher GM-ITE scores compared to 0 self-study time. Working 60–79 hours per week was predictive of a higher GM-ITE score (adjusted coefficient: 1.27; 95% confidence interval: 0.81-1.72; p<.001) compared to shorter working hours (<60 hours). The other factors significantly associated with GM-ITE scores were gender (women), participation in a general medicine rotation, internal medicine rotation duration, number of night shifts, and number of assigned patients.

## DISCUSSION

Resident physicians from training programs in regions with a low and moderate PUD index (areas facing physician shortages due to regional maldistribution) achieved significantly higher GM-ITE scores than those in programs with a high PUD index. To the best of our knowledge, this is the first study to quantitatively evaluate the relationship between the distribution of physicians and the core clinical competency of the resident physicians using nationwide data.

Physician maldistribution is a global problem that hinders patients’ ability to access healthcare services.^28^ Physicians are geographically maldistributed, with too few physicians in rural areas.^29,30^ In Japan, there is a well-documented issue of physician shortages and regional maldistribution, where approximately 90% of physicians are clustered in urban areas.^31^ As an example of countermeasures, changing the medical education system so that it selects, trains, and retains physicians who are more likely to practice in rural areas can remedy problems of rural geographic maldistribution.^29^

Resident physicians who train in areas with a high PUD index may have a more passive attitude towards clinical training than resident physicians who train in low PUD index areas. This could be attributed to the possibility that clinical experiences in areas with a high PUD index might not offer as diverse or challenging opportunities as those in areas with a lower PUD index. In high PUD index areas, opportunities for the supervising physician to allow resident physicians to make independent judgments may be limited, leading to an observational training style. Furthermore, the higher patient load or complexity in these high PUD index areas could potentially overshadow the emphasis on hands-on experience, prioritizing efficiency over educational value.^32^

Areas with high numbers of supervising physicians and nurses involved with resident physicians have disadvantages. When resident physicians interact with several different senior physicians and nurses, it takes time to establish relationships and build trust, which may lead to inefficient learning and reduced effectiveness. A lower number of supervising physicians in low PUD areas, including medical resident physicians, is associated with higher GM-ITE scores.^33^

Resident physicians in Japan require at least 60–65[duty hours per week during their clinical training to acquire a certain level of clinical knowledge.^21^ However, clinical knowledge does not proportionately increase with increases in duty hours. The optimal number of duty hours that maximizes the development of resident physicians’ competencies must be determined. In Japanese areas with a low PUD index, resident physicians face moderately longer working hours than resident physicians in areas with a high PUD index, which may contribute to skill development.

Resident physicians from areas with a low PUD index dedicated more time to self-study than those in areas with a high PUD index. Self-study time was strongly correlated with GM-ITE scores, suggesting that self-directed learning is crucial for enhancing competency, irrespective of the region (rural or urban). The positive relationship between longer self-study time and higher GM-ITE scores in this study is consistent with the results of a previous report.^34^ William Osler noted that, “To study the phenomena of disease without books is to sail an uncharted sea, while to study books without patients is not to go to sea at all.”^35^ Hence, it becomes imperative to ensure the optimal equilibrium between the two elements within the constraints of limited time. In Japan, physicians’ work-style reforms, including the establishment of working hour limits for physicians and resident physicians, are scheduled to be implemented in April 2024.^36^ Resident physicians working <60 hours per week reported less self-study time than those working 60–65 hours per week.^21^ Therefore, innovative self-study tools that efficiently demonstrate educational concepts in a limited time are needed.

Physician maldistribution remains a significant challenge in the global healthcare context. The findings of the current study underscore the importance of policy-driven interventions in addressing these disparities. The correlation between the PUD index and resident physicians’ clinical competence suggests that the quality of medical training and patient care is intricately linked to the equitable distribution of healthcare resources. The impact of physician distribution on clinical competence may have implications beyond Japan, suggesting a need for international collaborative efforts to address this issue.

### Limitations

This study is not without limitations. First, the potential for selection bias cannot be ruled out. As the GM-ITE is voluntary for training hospitals, only half of the resident physicians in Japan participated in this study. Second, a bias may exist between data from community and university hospitals. Similar to previous studies regarding the GM-ITE, resident physicians from university hospitals had lower GM-ITE scores in this study.^37^ The low PUD index group had a lower proportion of university hospitals, which may be a confounding factor. Third, the resident physicians’ baseline clinical skills were not assessed. The participants had various medical school experiences, which may have impacted the study results. Fourth, this investigation did not evaluate the actual number of supervising physicians in each hospital or their years of clinical experience. These factors may have affected the GM-ITE scores. Fifth, the current study relied heavily on the GM-ITE scores as a primary measure of clinical competence. Although the GM-ITE is a validated tool, it predominantly assesses theoretical knowledge rather than practical skills. Sixth, although the regions were categorized based on the PUD index, intra-regional disparities may exist. Different areas within a single region may have varying physician densities, which may lead to heterogeneity in the clinical exposure and training of residents within the same PUD category. Seventh, while the GM-ITE assesses medical interviewing for communication skills, it does not sufficiently evaluate crucial aspects such as interpersonal literacy and interprofessional communication essential for effective leadership in contemporary medicine.

## CONCLUSIONS

In prefectures with a shortage of physicians, resident physicians enrolled in training programs can attain favorable core clinical competency scores. This highlights the close relationship between efforts to address physician maldistribution and the enhancement of clinical competency among resident physicians in Japan. Regions with a relatively insufficient distribution of physicians, as indicated by a lower PUD index, have higher clinical proficiency, measured as the GM-ITE scores. As the healthcare landscape evolves, the assurance of equitable physician distribution has emerged as a central tenet of robust healthcare systems. The findings of this study have global implications.

## Article Information

### Authors’ contributions

Dr Kiyoshi Shikino and Dr Yuji Nishizaki had full access to all of the data in the study and take responsibility for the integrity of the data and the accuracy of the data analysis.

*Concept and design:* Kiyoshi Shikino, Yuji Nishizaki. *Acquisition, analysis, or interpretation of data:* All authors. *Drafting of the manuscript:* Kiyoshi Shikino.

*Critical review of the manuscript for important intellectual content:* All authors.

*Statistical analysis:* Koshi Kataoka, Masanori Nojima.

*Obtained funding:* Yuji Nishizaki.

Administrative, technical, or material support: Yuji Nishizaki, Yasuharu Tokuda.

Supervision: Yuji Nisizaki, Yasuharu Tokuda.

### Data Sharing Statement

See the Supplement 2.

### Additional Contributions

The authors express gratitude to JAMEP members and the members of the question development committee as well as the peer-review committee of the GM-ITE.

## Data Availability

All data produced in the present study are available upon reasonable request to the authors

## Supplement

**Supplement 1.**
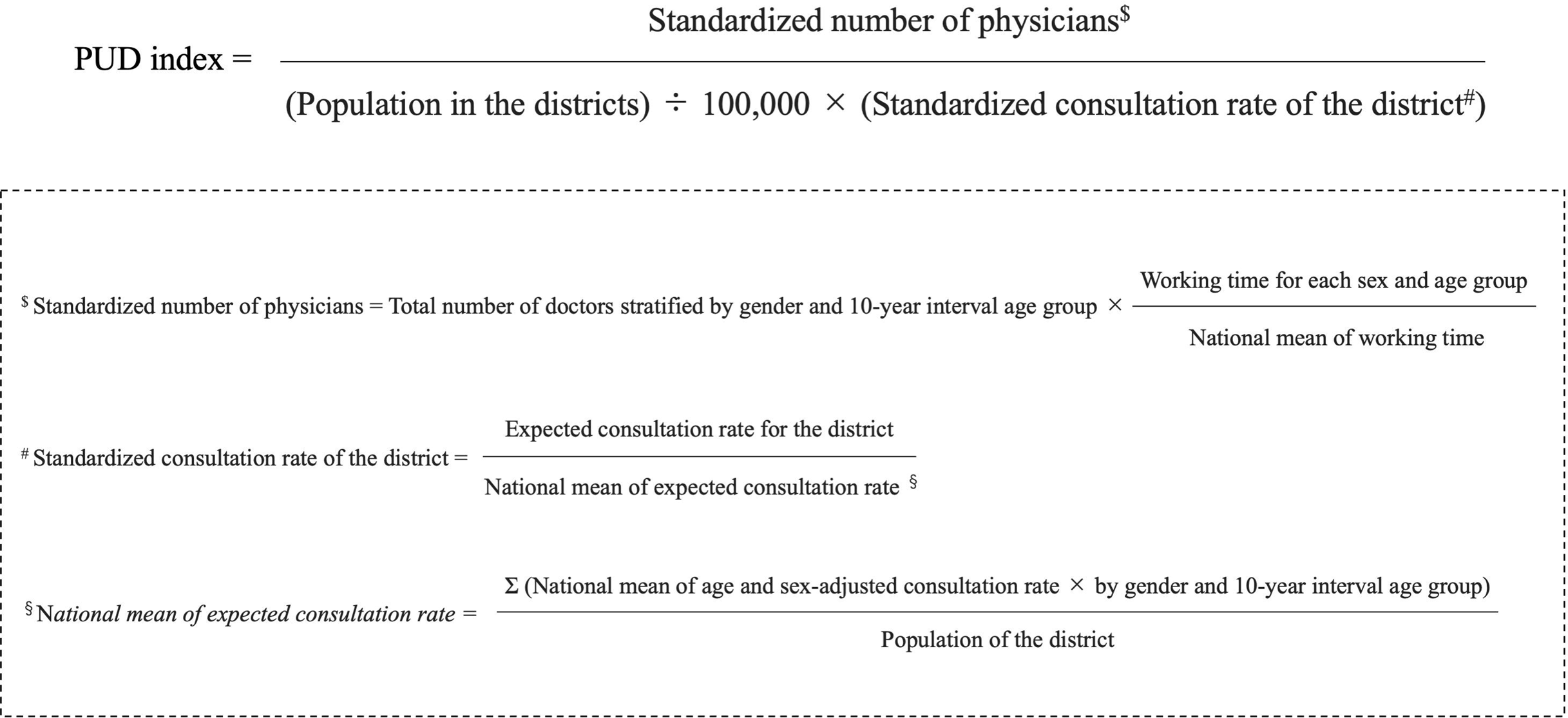
Physician uneven distribution index (PUD index)

Supplement 2. Data sharing statement.

## REFERENCES

1. Takayama A, Poudyal H. Incorporating medical supply and demand into the index of physician maldistribution improves the sensitivity to healthcare outcomes. J Clin Med. 2021;11(1):155.

2. Hara K, Kunisawa S, Sasaki N, Imanaka Y. Examining changes in the equity of physician distribution in Japan: A specialty-specific longitudinal study. BMJ Open. 2018;8(1):e018538.

3. Kobayashi Y, Takaki H. Geographic distribution of physicians in Japan. Lancet. 1992;340(8832):1391-1393.

4. Toyabe S. Trend in geographic distribution of physicians in Japan. Int J Equity Health. 2009;8:5.

5. Tanihara S, Kobayashi Y, Une H, et al. Urbanization and physician maldistribution: a longitudinal study in Japan. BMC Health Serv Res. 2011;11:260.

6. Wu J. Measuring inequalities in the demographical and geographical distribution of physicians in China: Generalist versus specialist. Int J Health Plan Manag. 2018;33(4):860–879.

7. Karan A, Negandhi H, Nair R, Sharma A, Tiwari R, Zodpey S. Size, composition and distribution of human resource for health in India: New estimates using National Sample Survey and Registry data. BMJ Open. 2019;9(4):e025979.

8. Morris S, Sutton M, Gravelle H. Inequity and inequality in the use of health care in England: An empirical investigation. Soc Sci Med. 2005;60(6):1251–1266.

9. Guttmann A, Shipman SA, Lam K, Goodman DC, Stukel TA. Primary care physician supply and children’s health care use, access, and outcomes: Findings from Canada. Pediatrics. 2010;125(6):1119–1126.

10. Mick SS, Lee SY, Wodchis WP. Variations in geographical distribution of foreign and domestically trained physicians in the United States: ‘safety nets’ or ‘surplus exacerbation’? Soc Sci Med. 2000;50(2):185–202.

11. Winkelmann J, Muench U, Maier CB. Time trends in the regional distribution of physicians, nurses and midwives in Europe. BMC Health Serv Res. 2020;20(1):937.

12. Organisation for Economic Co-operation and Development (OECD). Doctors. Paris: OECD Publishing. https://www.oecd-ilibrary.org/content/data/4355e1ec-en (2018, accessed 25 October 2023).

13. Sato H. Demand, supply and shortages of physicians: a critical analysis on the current government’s method. J Health Welf Policy. 2020;3:39–48. [Google Scholar]

14. Ministry of Health, Labour and Welfare Study Group on Supply and Demand of Medical Staff: Doctor Supply and Demand Subcommittee 4th Interim Report. [(accessed on 25 October 2023)];2019 Available online: https://www.mhlw.go.jp/content/12601000/000504403.pdf

15. Tago M, Shikino K, Hirata R, et al. General medicine departments of Japanese universities contribute to medical education in clinical settings: a descriptive questionnaire study. Int J Gen Med. 2022;15:5785–5793.

16. Teo A. The current state of medical education in Japan: a system under reform. Med Educ. 2007;41(3):302–308.

17. Kozu T. Medical education in Japan. Acad Med J Assoc Am Med Coll. 2006;81(12):1069–1075.

18. Ohde S, Deshpande GA, Takahashi O, Fukui T. Differences in residents’ self-reported confidence and case experience between two post-graduate rotation curricula: results of a nationwide survey in Japan. BMC Med Educ. 2014;14:141.

19. Ministry of Health, Labour and Welfare, Medical Practice Council, Committee for medical practitioner specialized training: Initiatives to enhance clinical training opportunities in the region. [(accessed on 25 October 2023)];2023 Available online: https://www.mhlw.go.jp/content/10803000/001152774.pdf

20. Fukui S, Shikino K, Nishizaki Y, et al. Association between regional quota program in medical schools and practical clinical competency based on General Medicine In-Training Examination score: a nationwide cross-sectional study of resident physicians in Japan. Postgrad Med J. 2023:99(1177):1197-1204.

21. Nagasaki K, Nishizaki Y, Shinozaki T, et al. Impact of the resident duty hours on in-training examination score: A nationwide study in Japan. Med Teach. 2022;44(4):433–440.

22. Nagasaki K, Nishizaki Y, Shinozaki T, Kobayashi H, Tokuda Y. Association between resident duty hours and self-study time among postgraduate medical residents in Japan. JAMA Netw Open. 2021;4(3):e210782.

23. Garibaldi RA, Subhiyah R, Moore ME, Waxman H. The in-training examination in internal medicine: an analysis of resident performance over time. Ann Intern Med. 2002;137(6):505–510.

24. Kinoshita K, Tsugawa Y, Shimizu T, et al. Impact of inpatient caseload, emergency department duties, and online learning resource on general medicine in-training examination scores in Japan. Int J Gen Med. 2015;8:355–360.

25. Shimizu T, Tsugawa Y, Tanoue Y, et al. The hospital educational environment and performance of residents in the general medicine in-training examination: a multicenter study in Japan. Int J Gen Med. 2013;6:637–640.

26. Nagasaki K, Nishizaki Y, Nojima M, et al. Validation of the general medicine in-training examination using the professional and linguistic assessments board examination among postgraduate residents in Japan. Int J Gen Med. 2021;14:6487–6495.

27. Tiffin PA, Illing J, Kasim AS, McLachlan JC. Annual Review of Competence Progression (ARCP) performance of doctors who passed Professional and Linguistic Assessments Board (PLAB) tests compared with UK medical graduates: national data linkage study. BMJ. 2014;348:g2622.

28. Elma A, Nasser M, Yang L, Chang I, Bakker D, Grierson L. Medical education interventions influencing physician distribution into underserved communities: a scoping review. Hum Resour Health. 2022;20(1):31.

29. Rosenthal MB, Zaslavsky A, Newhouse JP. The geographic distribution of physicians revisited. Health Serv Res. 2005;40(6 Pt 1):1931-1952.

30. Rosenblatt RA, Hart LG. Physicians and rural America. West J Med. 2000;173(5):348–351.

31. Ishikawa T, Nakao Y, Fujiwara K, Suzuki T, Tsuji S, Ogasawara K. Forecasting maldistribution of human resources for healthcare and patients in Japan: a utilization-based approach. BMC Health Serv Res. 2019;19(1):653.

32. Russell DJ, Wilkinson E, Petterson S, Chen C, Bazemore A. Family medicine residencies: How rural training exposure in GME is associated with subsequent rural practice. J Grad Med Educ. 2022;14(4):441–450

33. Mizuno A, Tsugawa Y, Shimizu T, et al. The impact of the hospital volume on the performance of residents on the General Medicine In-Training Examination: A multicenter study in Japan. Intern Med. 2016;55(12):1553–1558.

34. Nishizaki Y, Shimizu T, Shinozaki T, et al. Impact of general medicine rotation training on the in-training examination scores of 11, 244 Japanese resident physicians: a Nationwide multi-center cross-sectional study. BMC Med Educ. 2020;20(1):426.

35. Osler W. Address on the dedication of the new building. Boston Med Surg J. 1901;144:60–61.

36. Ministry of Health, Labour and Welfare. On the Revision of Overtime Regulations. [(accessed on 25 October 2023)];2019 Available online: https://www.mhlw.go.jp/content/10800000/000481338.pdf

37. Nishizaki Y, Nozawa K, Shinozaki T, et al. Difference in the general medicine in-training examination score between community-based hospitals and university hospitals: a cross-sectional study based on 15,188 Japanese resident physicians. BMC Med Educ. 2021;21(1):214.

